# Unified Measures Quantifying Intensity and Similarity of Pain and Somatosensory Percepts

**DOI:** 10.1101/2025.01.16.25320030

**Authors:** Eric J. Earley, Malin Ramne, Johan Wessberg

## Abstract

Across somatosensory and pain literature, there exist several methods of characterizing the location and extent of perceived sensations, and quantifying how these sensory maps may differ. However, these measures of somatosensory intensity and similarity can give non-unique results, creating challenges in literature review and meta-analysis across different methods. In this paper, we propose novel and unifying measures to quantify the similarity and intensity of pain maps and somatosensory percepts. These measures are generalizable and can be applied to any application of somatosensory maps, and are usable with both discretized and free-hand drawings in both 2D and 3D representations. Somatosensory Percept Intensity (SPI) is inspired by Piper’s Law, which describes the phenomenon of incomplete spatial summation wherein changes in pain area do not yield linearly proportional changes in perceived intensity. Somatosensory Percept Deviation (SPD) is derived from optimal transport theory, which quantifies differences between two probability distributions or somatosensory maps. Mathematical derivations for both measures are provided. The utility of these measures is demonstrated using data from two studies – one characterizing elicited somatosensory percepts, and one investigating neuropathic pain drawings. The proposed measures strongly agree with the validation studies, illustrating their potential as agnostic measures for characterizing somatosensory percepts in studies and meta-analyses. Ultimately, our work yields powerful unified measures for use in the fields of perception and pain, and may aid in improved pain characterization within healthcare, granting a better understanding of the needs and progression of patients experiencing pain.

**NEW & NOTEWORTHY:** We propose novel and unifying measures to quantify the similarity and intensity of pain maps and somatosensory percepts. These measures are generalizable and applicable to any application of somatosensory maps, resulting in improved pain characterization within healthcare and better understanding of the needs and progression of patients experiencing pain.

## INTRODUCTION

The mapping of localized sensations felt on the body is of significant interest in both clinical and scientific circles. Pain maps are commonplace in clinical evaluation and treatment of chronic pain such as fibromyalgia, spinal cord injury, and low back pain (1–6). In neuroscience research, advancements in peripheral nerve stimulation have led to many studies characterizing sensations elicited from such stimulation (7–11). In both cases, these somatosensory maps can localize nociceptive or non-nociceptive sensations via “free-hand” drawings or by indicating distinct regions on the body, which may be further discretized into subregions or uniform grids (**Figure 1**). The variety of maps results in a corresponding variety of methods to quantify “similarity”; regions or grids may be compared using the number of stable sites (9), whereas similarity between free-hand drawings may use image processing algorithms (12, 13) or the Jaccard similarity coefficient (5, 14–16). This variety can make comparison between studies difficult, even those which evaluate the same sensations.

**Figure 1:**
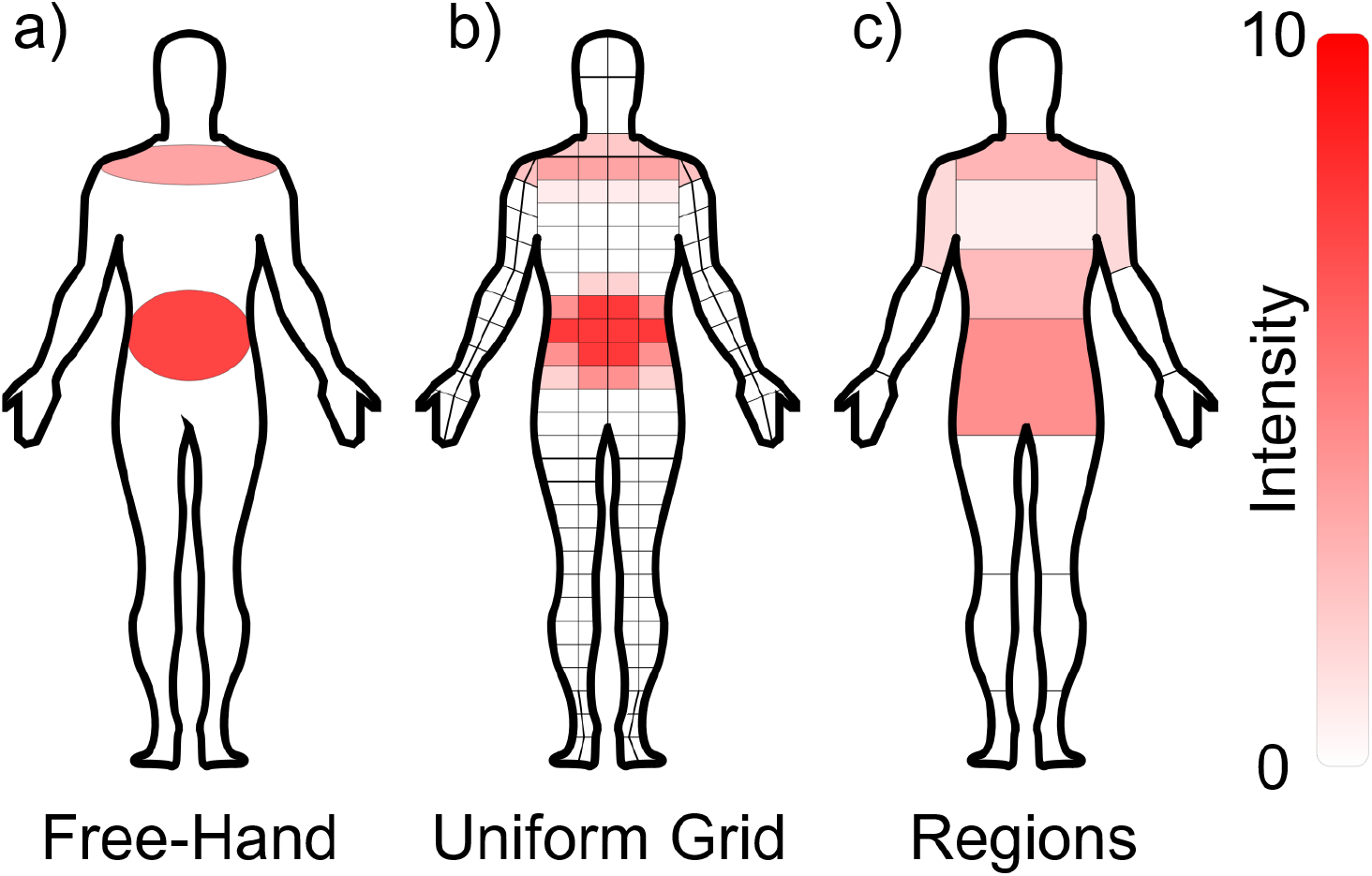
Pain maps and somatotopic maps can take on many forms, which can make cross-study comparisons challenging. Using a hypothetical instance of an individual indicating shoulder and back pain, we illustrate the differences between somatotopic maps recorded as free-hand drawings (a), a uniform grid (b), and body regions (c). In all instances, the sensible area is considered to be the extent of the body map, whereas the perceptive field is considered to be all areas where sensation is currently felt, with a corresponding intensity provided for each element of the perceptive field. The goal of the proposed measures is to describe differences between two perceptive fields in such a way as to consider both intensity and location, and such that results can be compared irrespective of recording method.

The intensity of pain or sensations also feature a wide variety of quantitative metrics. For pain especially, these metrics tend to be acquired via questionnaire, typically employing a Numeric Rating Scale (NRS) (17, 18). However, these measures alone are unable to decouple between the spread and the intensity of somatosensory percepts or quantify the change of pain intensity in different regions of the affected area in the context of pain. Hence, a measure for precisely quantifying intensity of sensations could be helpful for tracking progress and outcomes of pain interventions, as patients often struggle to articulate the nuances of their pain experience (19). Having a quantifiable measure of pain intensity beyond the NRS may allow for clearer communication between patients and healthcare providers, leading to better-informed decisions and care. Measures of pain intensity and spatial spread can also generate quantifiable feedback on the outcome of treatment decisions and can help optimize treatment plans and reduce unnecessary procedures or medications, potentially saving costs associated with pain management while improving outcomes for patients.

Another limitation of existing measures of similarity and intensity is that their behavior can be inconsistent in certain conditions. For example, measuring the similarity as the number of stable sites (9) will calculate less similarity if two percept fields are farther apart, but only if there is at least some overlap. If the fields share no common areas within the somatosensory map, then fields located close to each other will yield the same outcome as fields that are farther away. This non-monotonicity can, in some applications, complicate the interpretation of analyses made using these measures, thus there is a need for a new set of measures with consistent behavior across conditions.

In this manuscript, we propose novel and unifying measures to quantify intensity and similarity of somatosensory percepts, termed Somatosensory Percept Intensity (SPI) and Somatosensory Percept Deviation (SPD), respectively. These measures are generalizable and can be applied to any application of somatosensory maps, and are usable with both discretized and free-hand drawings in both 2D and 3D representations. We demonstrate the utility of these measures using data from two separate studies investigating somatosensation and pain using different methods.

## MATERIALS AND METHODS

The methods described below originally arose during the course of research characterizing somatosensory percepts elicited from direct nerve stimulation in persons with upper limb amputation who had been fitted with a neuromusculoskeletal prosthesis (20–22). The process of capturing qualitative descriptions of sensations, as well as understanding their spatial and temporal dynamics, involved frequent and direct involvement with participants, whose input has subsequently guided the somatosensory map framework and measures of intensity and similarity in the following sections.

### Assumptions about Somatosensory Maps

#### Percept Field

We define a percept field as an indication of the regions of the body on which a sensation is felt. Such a percept field is intrinsically bound to what we term the sensible area – the areas of the body capable of sensation in even the slightest degree, or the corresponding phantom representation of a missing portion of the body.

For the calculation of SPI and SPD, we make the following assumptions about percept fields:

1. The size of a percept field is greater than zero (e.g. a positive value) in all dimensions of interest.
2. The distance between any portion of any two percept fields can be measured or estimated in absolute (positive) units of length.
  a. For pre-defined grids and regions, distance (e.g. Euclidean norm) will typically be measured between region centroids or anchor points.
  b. For free-hand drawn boundaries, the distance between any two points between fields should be measurable.
3. The percept field has a discrete and closed boundary which may be defined by a combination of free-hand drawn boundaries, pre-defined grids and regions, and the limits of the corporeal body and phantom representation.
4. Any perceived sensation is an instantaneous capture of the percept intensity within the percept field and does not account for temporal properties such as transient changes in location or sensation area.

By virtue of assumptions (1) and (2), percept fields exist within a space where measurements can be described in terms of units of length including meters, pixels, or percentage body height. Assumption (3) bounds percept fields to the sensible area but also prohibits “feathered” borders as are collected via some questionnaires (23); a method to incorporate this “feathered” edge into the SPD measure is described in the **Appendix**. Assumption (4) limits sensation maps to instantaneous sensations, which allows the quantification of the acute variability and similarity of sensations.

#### Percept Intensity

We define the percept intensity as the magnitude of a perceived sensation. Such intensity is intrinsically bound to the percept field – all elements of the percept field *must* be accompanied by a percept intensity.

For the calculation of SPI, we make the following assumptions about percept intensity:

5. The percept intensity is greater than or equal to zero for all intensities.
6. The working range of percept intensity is finite.
7. The measure of percept intensity is monotonic across its working range.

Assumptions (5) and (6) ensure that percept intensity can be properly quantified and, along with (7), ensure the monotonicity of the SPI. Assumption (7), importantly, does not enforce a particular scaling, such as a linear relationship, and allows the measure to be described in terms of another measured unit, or to be a unitless value such as from a Likert-type scale.

### Desired Behavior of Similarity and Intensity Measures

Below, we list the desirable behavior of SPI:

1. Minimum intensity is achieved if there is no perceivable sensation within the region of interest (RoI)
2. Maximum intensity is achieved if maximum sensation is perceived throughout the entire RoI
3. A larger change in percept intensity yields a larger change in SPI when percept field area is constant
4. A larger change in percept field area yields a larger change in SPI when percept intensity is constant

Behaviors (1) and (2) define the minimum and maximum limits of percept intensity, whereas behaviors (3) and (4) align the directionality and ensure monotonicity of the intensity measure. Here, we list the desirable behavior of a measure of SPD:

5. Maximum similarity is achieved if two somatosensory maps are identical
6. A larger change in percept field area yields a larger reduction in similarity
7. A larger change in percept field position yields a larger reduction in similarity

Behavior (5) defines the concept of similarity as the degree of agreement between two maps, whereas behaviors (6) and (7) align the directionality and ensure monotonicity of the similarity measure.

### Derivation of Measures

#### Percept Map

Let ***M*** ∈ ℝ^*N*^ represent a percept map existing within an *N*-dimensional somatosensory map (e.g. a 2D drawing or a 3D body model). ***M*** can represent a collection of discrete points (as might be obtained from a somatotopic map divided into regions) or a continuous closed contour (as might be obtained from a free-hand drawn percept field). However, the method used to calculate SPD (defined in the next section) is not analytically tractable in most cases (though a generalization of this approach for continuous regions is described in the **Appendix**), thus continuous closed contours should be discretized into a finite number of elements so that the metric can be calculated via numerical optimization. We therefore let ***M*** comprise a total of *n*individual regions *M*(*i*), each encompassing an *N*-dimensional area *A*(*i*) and with a location described by a single coordinate ***x***_*i*_ ∈ ℝ^*N*^.

#### Percept Field

Let ***P***∈ ***M*** represent a measured percept field describing a somatosensory or pain experience. Each region of percept field *P*(*i*) is defined as the instantaneous sensation intensity for which 0 ≤ *P*(*i*) ≤ *P*_*max*_(*i*), where *P*_*max*_(*i*) is the maximum intensity that can be felt in that region; for regions outside of the somatotopic map or regions that are completely desensitized, *P*_*max*_(*i*) = 0, otherwise *P*_*max*_(*i*) will depend on the limits of the intensity scale used (e.g. 10/10 on a Likert scale) and the level of desensitization in the region.

#### Somatosensory Percept Intensity (SPI)

SPI encompasses the total “amount” of sensation perceived by a person. It can be thought of as a weighted sum of the quantified sensation intensities (e.g. as captured by an NRS) and the areas that each sensation is felt across (**Figure 2a**). The calculation of SPI is inspired by Piper’s Law, which describes spatial and temporal summation of luminance thresholds in peripheral vision (24, 25). The same patterns of spatial and temporal summation have also been noted in pain evoked by mechanical pressure (26). One phenomenon which arises from this prior research is the notion of incomplete summation – where changes in stimulus area do not result in a linearly proportional change in perceived intensity – which has been observed in vision (27–29). It is possible that the same phenomenon occurs in somatosensory stimuli, though to our knowledge no study has yet investigated this.

**Figure 2:**
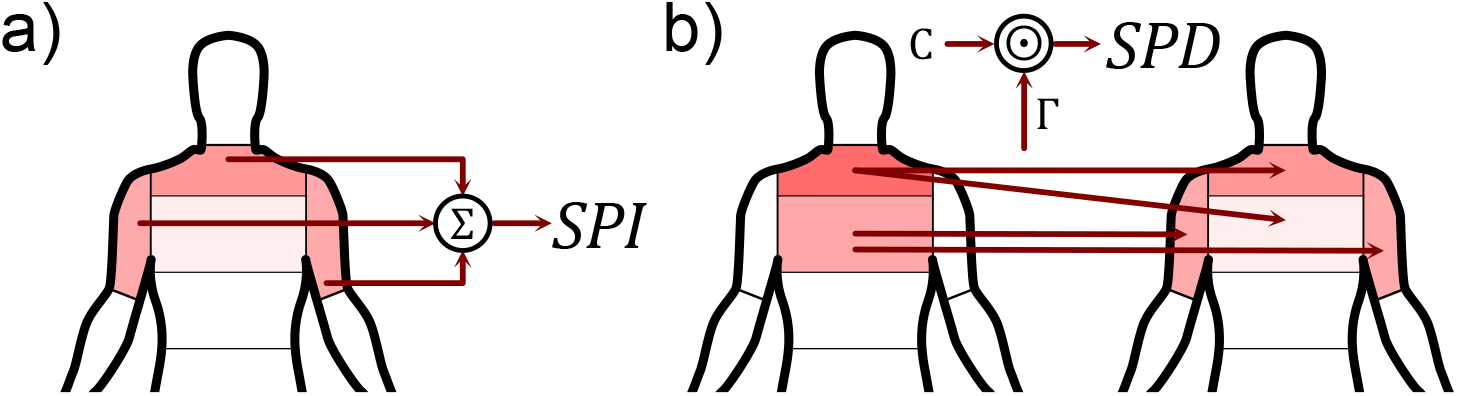
Intensity and similarity measures presented here can be calculated from somatotopic and pain maps. (a) Somatosensory Percept Intensity (SPI) is a weighted sum of the intensity of a sensation in a region and the size of that region. (b) Somatosensory Percept Deviation (SPD) is the element-wise product (⊙) of the transportation plan ***Γ*** (describing the percept intensity changes from one percept field to another) and the transportation cost ***C***.

We calculate SPI as:

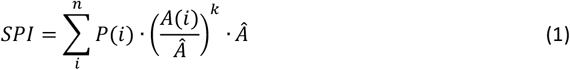

where *Â* is the unit area. Recall that *P*(*i*) and *A*(*i*) are the instantaneous sensation intensity and area of the *i*^*th*^ region of the percept map ***M***. We include a summation coefficient *k* in (1) to account for the possibility of incomplete summation. The summation coefficient can range from 0 to 1, asserting different assumptions upon the system or simplifying calculations. A summation coefficient of 0, for example, may be used if all regions are of equal size (or otherwise carry equal weight) to effectively remove *A*(*i*) from the equation, whereas a summation coefficient of 1 assumes complete spatial summation. As summation coefficients may vary widely between applications, we recommend the default value of *k* = 0.5 commonly cited in literature (24, 25) unless prior research suggests the use of a different coefficient.

The units of SPI are the product of the units of the sensation intensity and the area. These units allow for intuitive interpretation of the outcome within the context of its measurement, however it can make comparison difficult between studies using different measures of intensity. In such instances where comparison is desired, the normalized SPI (nSPI) can be calculated as:

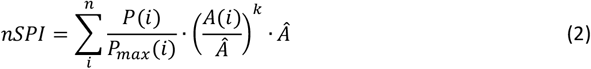

Defining SPI and nSPI in this way ensures that total experienced sensation scales according to both instantaneous sensation intensity and percept field area. nSPI also ensures parity between studies using different scales and allows for the use of biological markers as a quantitative measure of intensity (e.g. afferent nerve or cortical activity magnitudes).

#### Somatosensory Percept Deviation (SPD)

Given two somatosensory maps (e.g. two pain maps recorded at different times), SPD quantifies the degree of difference between the maps. The amount of deviation between the two maps is, by definition, inversely related to the similarity of the maps (**Figure 2b**). To ensure compliance with the aforementioned assumptions and behaviors, SPD is based on Optimal Transport Theory (OTT), which strives to quantify “distance” between two probability distributions. In this case, we define the “probability distribution” as the indicated percept field, and thus OTT quantifies the “distance” between two percept fields. Specifically, SPD uses the Wasserstein metric to quantify the dissimilarity between two percept fields (30, 31); this metric has seen similar use to explain dissimilarity between stochastic neural representations (32) and to classify prosthetic limb movements by comparing high-density EMG images to determine EMG maps with the greatest degree of similarity (33).

For SPD, the objective of OTT is to minimize the “cost” of transforming one percept field ***P***_**1**_ into another ***P***_**2**_. The transportation cost ***C*** is typically simply the Euclidean distance between regions *i* and *j*:

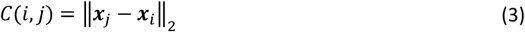

This Euclidean distance can be defined in any units of length (e.g. millimeters or pixels), however to facilitate the greatest degree of comparison between applications, we recommend normalizing the distance by using the 50^th^-percentile dimensions of the RoI, defined in anthropometric tables (34, 35), as a normalizing factor by which to divide the measured lengths of the somatotopic map.

The process of transforming ***P***_**1**_ into ***P***_**2**_, each comprising *n*regions, can be defined as the transportation plan ***Γ***, which takes the form of an *n*x *n*matrix where each element *Γ*(*i, j*) describes the quantity transferred from ***x***_*i*_ to ***x***_*j*_. For ***Γ*** to be valid for OTT, it must satisfy the following constraints:

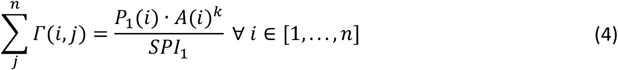

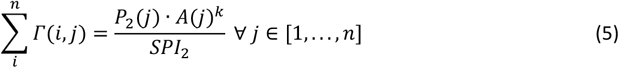

The normalizations by *SPS*_1_ and *SPS*_2_ ensure that the total transported quantity is equivalent. This normalization can also be achieved by defining the normalized percept field as:

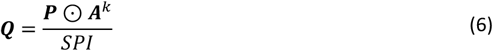

where ⊙ denotes the element-wise product. Applying (6) to (4)-(5) yields the following constraint equations:

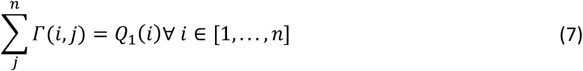

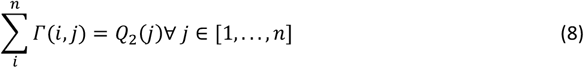

These constraints ensure that the total quantity transported out of *Q*_1_(*i*) is equal to the quantity contained within *Q*_1_(*i*), and that the total quantity transported into *Q*_2_(*j*) is equal to the quantity contained within *Q*_2_(*j*).

The final constraint pertains to the transportation plan ***Γ***, which cannot include negative values:

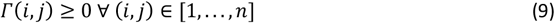

The total cost of executing transportation plan ***Γ*** can be calculated via element-wise product with the transportation cost ***C***. Formatting this as an optimization problem thus takes the form:

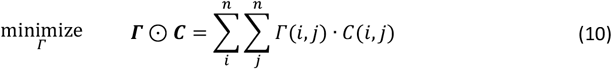

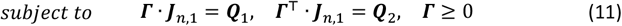

where ***J***_*n*,1_ is an *n*× 1 matrix of ones.

The estimated optimal transportation plan 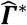 is then used to calculate SPD:

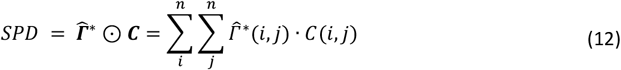

A smaller SPD outcome is associated with a greater degree of similarity between the somatotopic maps for percept fields ***P***_**1**_ and ***P***_**2**_.

The term “deviation” is also appropriate in this context, as the unit of an SPD measure is the same as the units of the transportation cost ***C***. Furthermore, it can be shown that SPD of a purely translational difference between ***P***_**1**_ and ***P***_**2**_ is simply the magnitude of the translation. Although it was not listed as a required behavior for similarity measures, one desirable property of SPD is its symmetry (*SPS*(***P***_**1**_ → ***P***_**2**_) = *SPS*(***P***_**2**_ → ***P***_**1**_)).

### Validation of Measures

To demonstrate the use of SPD as a similarity measure, we applied SPD to somatosensory data from two different studies. These studies were selected to represent a variety of somatosensory map styles, bodily RoIs, subject populations, and research purposes. For each study, SPD is used to quantify similarity between somatosensory percepts, and subsequently compare this result to those reported in the original study.

#### Use of Stimulation Waveform Shapes to Elicit Different Somatosensory Percepts

In this study by Collu et al. (11), transcutaneous electrical nerve stimulation (TENS) of the median nerve elicited sensations on the palms of the hands of eleven able-bodied individuals. The shapes of the stimulation waveforms were changed between various current profiles, and the location of the elicited sensation was recorded using a custom graphical interface. This interface allowed participants to encircle the area on the palm that a sensation was felt, after which the application would fit a bivariate normal distribution (appearing as an ellipse) to the indicated points. Participants were also asked to describe the quality, intensity, diffuseness, depth, dynamics, and naturalness of the elicited sensation. Although intensity was not evaluated in the study, the change in perceptive field area was investigated, showing that perceptive fields elicited by non-rectangular waveforms generally grew in area compared to rectangular waveforms, though these differences were not statistically significant.

SPD is used to quantify the differences in sensation intensity and percept fields between the five stimulation waveform shapes included in the study. This analysis includes the intensity *P*(*i*) omitted from the original study. The original study was approved by the Swedish regional ethical committee in Gothenburg (Dnr: 2019–05,446) and the research was performed in accordance with the relevant guidelines and regulations in compliance with the Declaration of Helsinki.

#### Reliability of Pain Drawings of Spinal Cord Injury Neuropathic Pain

In this study by Rosner et al. (6), twenty individuals with spinal cord injury (SCI) indicated areas in which they feel neuropathic pain by drawing on a ventral/dorsal body chart printed on a sheet of paper. These drawings were then scanned and the body was segmented into regions using a dermatome grid.(2) This process was then repeated approximately two weeks later, and the two pain maps were compared using interclass correlation coefficients, showing good agreement for both pain extent and intensity between measurements. This test-retest reliability is reanalyzed using SPD to investigate the degree of agreement for measures unifying both pain extent and pain intensity. The original study was approved by the local ethics board, “Kantonale Ethikkommission Zürich” (reference number: EK-04/2006).

## RESULTS

### Comparison with Existing Methods

For somatosensory intensity (**Table 1**), both methods will fulfill behaviors (1) and (2), but the Likert scale does not fulfill behaviors (3) and (4). The Likert scale is independent from the location and size of the percept field, thus maximum intensity can be achieved regardless of the size of the percept field.

**Table 1:** Somatosensory Percept Intensity comparison to existing methods.

| Behavior | Likert Scale | Somatosensory Percept Intensity (SPI) |
| --- | --- | --- |
| 1. Minimum intensity is achieved if there is no perceivable sensation within the region of interest (RoI) | ✓ | ✓ |
| 2. Maximum intensity is achieved if maximum sensation is perceived throughout the entire RoI | ✗ | ✓ |
| 3. A larger change in percept intensity yields a larger change in overall intensity when percept field area is constant | ✓ | ✓ |
| 4. A larger change in percept field area yields a larger change in overall intensity when percept intensity is constant | ✗ | ✓ |

Behaviors guaranteed across all scenarios are indicated in blue, whereas behaviors which are only sometimes fulfilled are indicated in grey; behaviors which can never be fulfilled are indicated in red.

Furthermore, while it is likely that respondents may indicate a higher percept intensity using a Likert scale when the percept field is larger, this would likely be more closely related to ambiguous or imprecise instructions given to the respondent when reporting their perceived sensation, rather than a feature of the Likert scale itself as a reporting method. Only the SPI is guaranteed to fulfill all four behaviors.

For somatosensory map location and area (**Table 2**), behavior (5) is fulfilled by all five methods, but behavior (6) is not guaranteed when measuring the overlapping area or number of stable sites, as a larger percept field which completely overlaps a smaller field will have the same overlapping area regardless of the size difference. Behavior (7) can never be captured when only measuring the change in percept area, and is not guaranteed when measuring overlapping area, structural similarity index, or Jaccard similarity coefficient (specifically, when both sensory maps do not overlap). Only the SPD is guaranteed to fulfill all three behaviors.

**Table 2:** Somatosensory Percept Deviation comparison to existing methods.

| Behavior | Change in Area | Overlapping Area/Stable Sites | Structural Similarity Index | Jaccard Similarity Coefficient | Somatosensory Percept Deviation (SPD) |
| --- | --- | --- | --- | --- | --- |
| 5. Maximum similarity is achieved if two somatosensory maps are identical | ✓ | ✓ | ✓ | ✓ | ✓ |
| 6. A larger change in percept field area yields a larger reduction in similarity | ✓ | ? | ✓ | ✓ | ✓ |
| 7. A larger change in percept field position yields a larger reduction in similarity | X | ? | ? | ? | ✓ |

Behaviors guaranteed across all scenarios are indicated in blue, whereas behaviors which are only sometimes fulfilled are indicated in grey; behaviors which can never be fulfilled are indicated in red.

### SPD Validation

#### Stimulation Waveform Shapes

**Figure 3** shows the SPI for sensations elicited by single pulses or trains of pulses of different neurostimulation waveform shapes. Percept intensities tended to be lowest for Rectangular waveforms (which deliver the lowest current while maintaining the same overall charge injection) and highest for Triangular waveforms (which deliver the highest current) Although percept intensity was not investigated in the original study, these trends align with the lower rheobasic currents identified for non-rectangular waveforms; however, no significant differences in percept intensities were found between waveform shapes for either single pulses (*p* ≥ 0.0537) or for pulse trains (*p* ≥ 0.0537) of suprathreshold stimuli.

**Figure 3:**
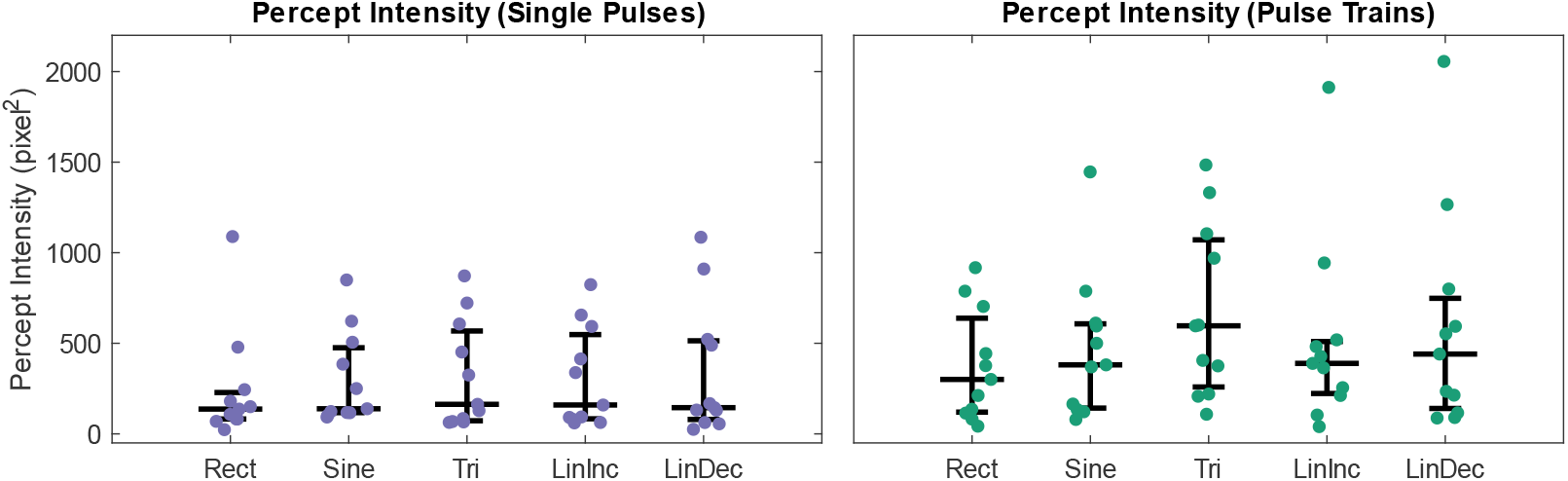
Percept Intensities for sensations elicited by different neurostimulation waveform shapes, delivered as either single pulses (lilac) or as trains of pulses (green).

**Figure 4** shows the SPD between somatosensory maps elicited by different neurostimulation waveform shapes. Somatosensory maps (inset) which were more dissimilar resulted in a higher percept deviation. For single pulses, percept deviations between Triangular and Linear Increasing waveforms were smaller than deviations between Rectangular and Triangular waveforms (*p* = 0.032), and deviations between Sinusoidal and Triangular waveforms (*p* = 0.042). For pulse trains, percept deviations between Rectangular and Triangular waveforms were significantly larger than deviations between Triangular and Linear waveforms (*p* ≤ 0.042). These results support the hypothesis that Triangular and Linear waveforms are more similar than Rectangular or Sinusoidal waveforms. However, most other comparisons were not significantly different.

**Figure 4:**
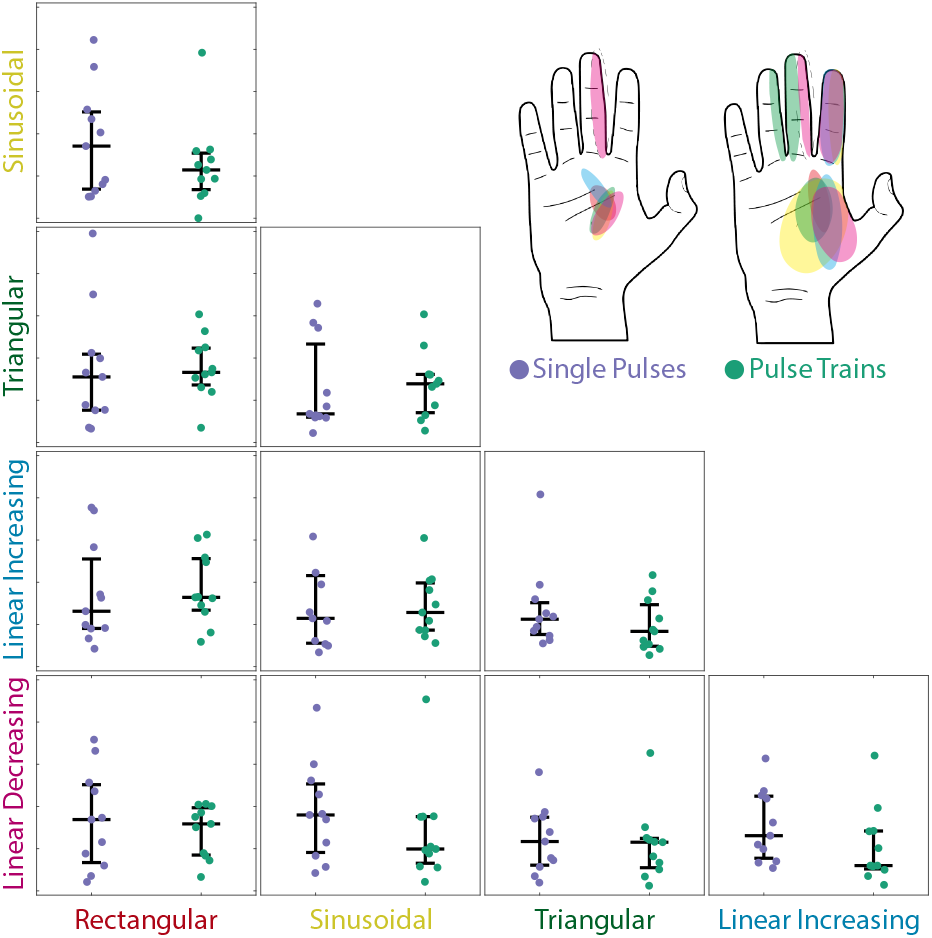
Percept Deviation between somatosensory maps elicited by different neurostimulation waveform shapes (inset, top-right), delivered as either single pulses (lilac) or as trains of pulses (green). Percept deviations range from 0 to 250 pixel-units in the plots shown.

#### Neuropathic Pain Drawings

**Figure 5** shows the outcomes of the proposed metrics in comparison to the outcomes reported in the original study. The scatter plots (left plots) show the mean neuropathic pain extent (e.g. the percentage of the body on which pain was indicated), mean neuropathic intensity (NRS scale 0-10), and the somatosensory Percept Intensity, and their relations to SPD. Across all three metrics, there appears to be no clear trend, suggesting independence of the SPD from pain extent and pain intensity separately, and from SPI (which, in essence, is jointly influenced by pain extent and intensity).

**Figure 5:**
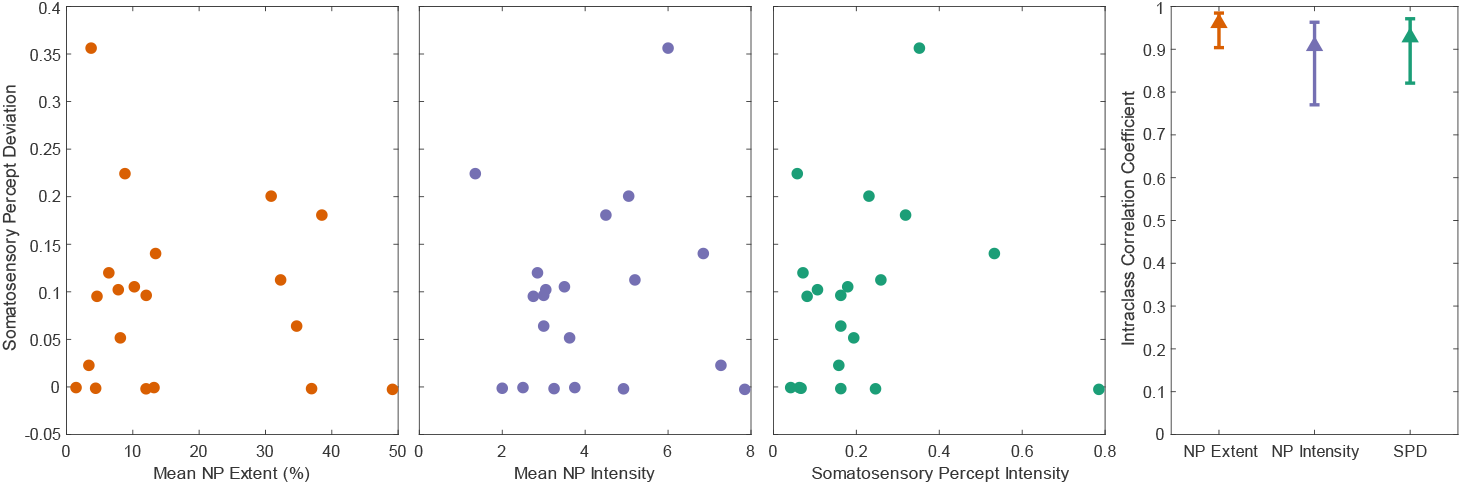
Scatter plots illustrate the general independence of the neuropathic pain (NP) extent and intensity, and the SPI, from the SPD (left plots). The Intraclass Correlation Coefficient (right plot) also showed no major differences between the metrics of NP extent, NP intensity, and SPD.

The right plot shows the Intraclass Correlation Coefficients (ICC) for pain extent and intensity (which are reported in the original study) and for SPD. All three metrics showcase a strong degree of ICC, indicating strong agreement between test and retest outcomes. In summary, the proposed measures agree strongly with the findings of the original study.

## DISCUSSION

Here, we propose a set of unifying measures which aim to quantify the similarity and intensity of somatosensory percepts. Our intent was to develop measures which are generalizable to the methods which are used to capture these somatosensory maps, be they discretized or continuous in nature (**Figure 1**). After defining the design criteria and deriving SPD and SPI, we validate the measures by reanalyzing data from two studies using two different methods of somatosensory mapping.

In the first study, percept fields were drawn to indicate the perceived sensations elicited from TENS of the median nerve in 11 subjects. The original study only compared perceptive field area between stimulation conditions, but by analyzing the same data using SPD we were able to quantify not only the differences in percept field area, but also location. The importance of this distinction can be seen when comparing the results of the original study to the results presented here. Percept intensities were, as expected, higher for stimulation pulse trains than for single pulses (**Figure 3**); however, the deviation between percept fields was smaller for pulse trains. This suggests that the percept fields for single pulses were less stable between stimulation conditions, which may be due to percept field saturation for pulse trains.

In the second study, locations of neuropathic pain were recorded from twenty individuals twice, and the percept fields were segmented into regions using a dermatome grid. The size of the percept field was quantified as the extent of the marked areas, as a percentage of the total area, and intensity was measured with a Likert scale. Measuring repeatability of neuropathic pain using SPD yielded a similar intraclass correlation coefficient to using the original measures, overall demonstrating agreement with the original study.

In studies, pain is often reduced to one or a handful of numbers as the proxy for the intensity and unpleasantness of certain aspects of the pain. In reality, pain is a complex and highly subjective experience, encompassing myriad sensation qualities. Furthermore, pain resulting from and injury of lesion can spread to secondary locations through processes of peripheral and central sensation (36). The quality and intensity of such “referred” pain can differ significantly from the local pain at the location of the injury, and importantly the underlying mechanism of the referred pain can be separate from the cause of local pain (37). Different pain interventions target different mechanisms, and thus referred and local pain may be differentially affected (36). Evaluating pain as a single point on an NRS may not capture changes in pain intensity at different sites, and thus may not accurately reflect the change in sensation. Incorporating this type of measure in future pain studies could provide insight into the differential mechanisms of different pain sensations and treatments.

Another application of this measure could be in the realm of sensory phenotyping of pain, in which researchers and clinicians try to match sensory symptoms to pain mechanisms to better predict treatment responses of individual patients (38, 39). These sensory symptoms include, among other things, the intensity of different sensation qualities and how the intensity varies over time. SPD could add more nuance and information to this type of measure by also incorporating how the spatial spread of pain varies, and thus might allow for more accurate predictions of pain mechanism and treatment outcome.

In summary, we demonstrate that outcomes calculated using SPI and SPD are in agreement with the measures used in these different studies and can even provide new insights not available through other measures (as was the case with the neurostimulation study). Furthermore, SPI and SPD exhibit all of the desired behaviors listed in Tables 1 and 2. Thus, we believe SPI and SPD to be useful measures for studies involving the quantification of somatosensory percepts and how they may differ between conditions, providing unifying metrics comprising somatosensory percept intensity and deviation.

## Limitations

SPD has a computational complexity of up to *O*(*n*^3^ *log n*) (40), which can be prohibitively costly for somatosensory maps with many regions. The use of techniques such as entropic regularization has been proven to lower the computational complexity to nearly *O*(*n*) (41). Furthermore, the scope of the optimization can be reduced by removing regions *P* not contained in either ***P***_**1**_ or ***P***_**2**_.

The use of Likert scales or other subjective quantitative scales of pain has well-known limitations, particularly when it comes to reliability of responses and accuracy of pain memory (42, 43). Despite these limitations, these pain assessment tools are prevalent throughout numerous pain pathologies and are frequently standard of care within medicine (44). Our unified pain measures were developed assuming these standardized clinical practices, despite the aforementioned limitations, however the derivation of SPI and SPD are intentionally non-prescriptive so that other future measures and instruments could be used instead. There is great interest in identifying objective measurements of pain. In research, neurologically-derived signals are being used to decode somatosensory intensity, for example resulting from tactile stimulus (45). Other techniques proposed in research studies to quantify somatosensory intensity include heart rate variability (46) and fMRI (47). Despite the numerous limitations and lack of validation preventing these approaches from being used in clinical practice, these types of objective measures could be used to encode changes in intensity arising from changes in sensory field for the purposes of calculating SPI and SPD. Furthermore, these techniques may not be as suitable for clinical practice owing to the requirement of additional, and sometimes expensive, sensors.

## Conclusions

In this study, we introduce two new measures, Somatosensory Percept Intensity (SPI) and Somatosensory Percept Deviation (SPD), to better quantify the intensity and similarity of somatosensory percepts. These measures overcome several limitations of existing methods and can be applied to both pain and somatosensory maps. SPI and SPD fulfill all desired behaviors for similarity and intensity metrics and offer a unified approach to evaluating somatosensory percepts, which can improve the characterization of nociceptive and non-nociceptive sensory experiences and clinical outcomes, ultimately serving as the basis to create powerful new tools for researchers and clinicians alike.

## APPENDIX

### Generalized Definition of Somatosensory Percept Variability

Let *M* ∈ ℝ^*N*^ represent a percept field existing within an *N*-dimensional somatosensory map. Each region *M*(*x*) is defined as the instantaneous sensation intensity *P*(*x*) felt at a region with *N*-dimensional area *A*(*x*), for which 0 ≤ *P*(*x*) ≤ *P*_*max*_(*x*), where: *P*_*max*_(*x*) is the maximum intensity that can be felt in that region; for regions outside of the somatotopic map or regions that are completely desensitized, *P*_*max*_(*x*) = 0, otherwise *P*_*max*_(*x*) will depend on the level of desensitization in the region and the limits of the intensity scale used (e.g. 10/10 on a Likert scale).

#### Somatosensory Percept Intensity

We then calculate SPI as:

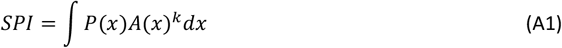

Defining SPI in this way ensures that total experienced sensation scales according to both instantaneous sensation intensity and percept field area.

#### Somatosensory Percept Deviation

Instead of a transportation plan ***Γ*** describing transport from one element to another, ***Γ*** is now a transportation field between percept fields ***P***_**1**_ and ***P***_**2**_. Calculating SPD now takes the form of the optimization problem:

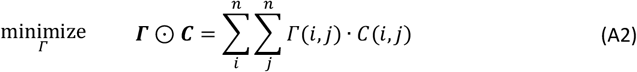

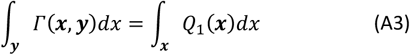

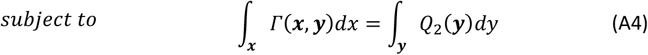

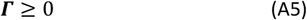

The optimal transport plan can once again be used to calculate SPD:

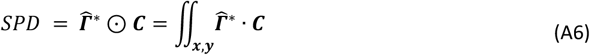

#### Extensions

There may be applications where a uniform transportation cost ***C*** is not desired when comparing somatosensory maps. For example, a study investigating selectivity of direct nerve stimulation to elicit tactile sensations may want to consider the mechanoreceptor density of the distal limb. In such a case, the transportation cost can be modified to better account for the variables of interest, for example by scaling the transportation cost inversely proportional to the two-point discrimination performance.

Sometimes, when defining the perceived locations of perceptions elicited from neurostimulation, the diffuseness of the sensation is also recorded, allowing for the subject to indicate whether the borders of the sensation are well-defined or “feathered”. This diffuseness could interfere with assumption (3), which states that percept fields must have a discrete border. However, the diffuseness of a sensation can be accounted for by introducing a diffuseness variable *β* to the definition of a modified percept field ***P****′*, such that:

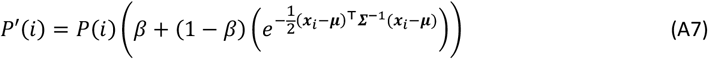

In other words, the modified sensation intensity is a diffuseness-weighted average between a uniform distribution (i.e., a Dirac measure) and a normalized bivariate Gaussian distribution, scaled by the original sensation intensity *P*(*i*). It should be noted that, although ***P****′* ≤ ***P***, SPD will remain unchanged due to the normalization in Equation 6.

## GLOSSARY

*M*: Percept map
***x***_*i*_: Coordinate of percept map region *M*(*i*)
*A*(*i*): Area of percept map region *M*(*i*)
*Â*: Unit area
*P*: Percept field of instantaneous sensation intensities within percept map ***M***
*Pmax*(*i*): Maximum sensation intensity of percept field region *P*(*i*)
*SPS*: Somatosensory Percept Intensity
*nSPS*: Normalized Somatosensory Percept Intensity
*k*: Summation coefficient
*SPS*: Somatosensory Percept Deviation
*P*: Percept field
*C*: Transportation cost
*Γ*: Transportation plan
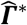: Estimated optimal transportation plan
*Q*: Normalized percept field
*J*: Matrix of ones
⊙: Element-wise product

## DATA AVAILABILITY

No original data were generated as part of this study. All data used in this study can be requested from the respective corresponding authors (6, 11).

## ACKNOWLEDGMENTS

We thank Jan Rosner, MD, and Michèle Hubli, PhD, for sharing their data of pain drawings for the validation performed in this study (6). We thank Riccardo Collu for his data using stimulation waveform shapes to elicit different somatosensory percepts via TENS (11). We thank Dr. Morten Bak Kristoffersen for his insights in developing the mathematics behind Somatosensory Percept Variability. EJE acknowledges Prof. Alex H Williams for his blog post, A Short Introduction to Optimal Transport and Wasserstein Distance (48).

## GRANTS

This work was supported by the Promobilia Foundation (Stiftelsen Promobilia), the IngaBritt and Arne Lundbergs Foundation (IngaBritt och Arne Lundbergs Forskningsstiftelse), and the Swedish Research Council (Vetenskapsrådet).

## DISCLOSURES

All authors declare that they have no conflicts of interest with the contents of this manuscript.

## AUTHOR CONTRIBUTIONS

EJE conceived and designed research, analyzed data, interpreted results, prepared figures, drafted manuscript, edited and revised manuscript, and approved final version of manuscript. ML and JW designed research, edited and revised manuscript, and approved final version of manuscript.

## Notes

### Competing Interest Statement

The authors have declared no competing interest.

### Author Declarations

De-identified somatosensory data from two different previous studies were used to validate the proposed measures. The study by Collu et al., 2023, was approved by the Swedish regional ethical committee in Gothenburg (Dnr: 2019-05,446) and the research was performed in accordance with the relevant guidelines and regulations in compliance with the Declaration of Helsinki. The study by Rosner et al., 2021, was approved by the local ethics board, "Kantonale Ethikkommission Zurich" (reference number: EK-04/2006).

